# Comparing facial feature extraction methods in the diagnosis of rare genetic syndromes

**DOI:** 10.1101/2022.08.26.22279217

**Authors:** Alexander J M Dingemans, Bert B A de Vries, Lisenka E L M Vissers, Marcel A J van Gerven, Max Hinne

**Affiliations:** Department of Human Genetics, Donders Institute for Brain, Cognition and Behaviour, Radboud University Medical Center, Geert Grooteplein Zuid 10, 6500 HB, P.O. Box 9101, Nijmegen, the Netherlands; Department of Artificial Intelligence, Donders Institute for Brain, Cognition and Behaviour, Radboud University, Thomas van Aquinostraat 4, 6525 GD Nijmegen, the Netherlands

**Keywords:** artificial intelligence, genetics, machine learning, facial recognition, deep learning

## Abstract

**Background and Objective:** Since several genetic disorders exhibit facial characteristics, facial recognition techniques can help clinicians in diagnosing patients. However, currently, there are no open-source models that are feasible for use in clinical practice, which makes clinical application of these methods dependent on proprietary software.

**Methods:** In this study, we therefore set out to compare three facial feature extraction methods when classifying 524 individuals with 18 different genetic disorders: two techniques based on convolutional neural networks (VGGFace2, OpenFace) and one method based on facial distances, calculated after detecting 468 landmarks. For every individual, all three methods are used to generate a feature vector of a facial image. These feature vectors are used as input to a Bayesian softmax classifier, to see which feature extraction method would generate the best results.

**Results:** Of the considered algorithms, VGGFace2 results in the best performance, as shown by its accuracy of 0.78 and significantly lowest loss. We inspect the features learned by VGGFace2 by generating activation maps and using Local Interpretable Model-agnostic Explanations, and confirm that the resulting predictors are interpretable and meaningful.

**Conclusions:** All in all, the classifier using the features extracted by VGGFace2 shows not only superior classification performance, but detects faces in almost all images that are processed, in seconds. By not retraining VGGFace2, but instead using the feature vector of the network with its pretrained weights, we avoid overfitting the model. We confirm that it is possible to classify individuals with a rare genetic disorder (thus by definition using a small dataset) using artificial intelligence and open-source all of the models used in this study, being the first study to open-source deep learning algorithms to be used to assess facial features in clinical genetics.

**Concise abstract:** Since several genetic disorders exhibit facial characteristics, facial recognition techniques can help clinicians in diagnosing patients. However, there are no open-source models available that are feasible for use in clinical practice, which makes clinical application of these methods dependent on proprietary software. This hinders not only use in clinic, but academic research and innovation as well. In this study, we therefore set out to compare three facial feature extraction methods for classifying 524 individuals with 18 different genetic disorders: two techniques based on convolutional neural networks and one method based on facial distances. For every individual, all three methods are used to generate a feature vector of a facial image, which is then used as input to a Bayesian softmax classifier, to compare classification performance. Of the considered algorithms, VGGFace2 results in the best performance, as shown by its accuracy of 0.78 and significantly lowest loss. We inspect the learned features and show that the resulting predictors are interpretable and meaningful. We confirm that it is possible to classify individuals with a rare genetic disorder (thus by definition using a small dataset) using artificial intelligence and open-source all of the models used in this study. This is the first study to open-source deep learning algorithms to assess facial features in clinical genetics.

## 1 Introduction

In the last decade, advances in computer vision have dramatically improved the available techniques for automated facial recognition. Since human-level accuracy is now possible with deep learning, convolutional neural networks (CNNs) are increasingly applied in our society — with applications ranging from face verification on airports to detecting facial emotions to gauge customer satisfaction [1, 2, 3].

Other applications of facial recognition using deep learning include the medical domain. In clinical genetics, for instance, these new techniques assist clinicians in recognizing genetic disorders in patients with neurodevelopmental disorders (NDD) [4, 5, 6, 7, 8, 9, 10]. This is possible since a significant portion of these patients have identifiable facial characteristics — Down’s syndrome is an example that most people will be able to recognize, but there are thousands of other genetic syndromes associated with specific and characteristic facial features. By harnessing the power of deep learning, we can train CNNs to distinguish between patients with different genetic disorders and, therefore, various facial features, lowering the possibility of a missed diagnosis. This ensures patients worldwide get access to the diagnosis they need for accurate surveillance and treatment — which due to the cost of genetic testing, is not a given.

Using these CNNs, we and others have shown remarkable results in recognizing genetic syndromes [5, 7, 10]. However, these applications use proprietary algorithms not available to the academic community, let alone the general population. In the context of open science, we think it is essential to open source these methods so that they can be verified and validated by the community and built upon and improved. While our previously published approach [6] uses open-source tools, use in the clinic is currently not feasible due to the significant time it takes to process a photograph, and because the model does not always recognize the face in an image. Furthermore, we hypothesize that the classification performance of this model can be improved if we employ the recent advances in facial recognition techniques.

However, while CNNs are powerful, they do not provide a clear-cut manner to inspect the models. This can be important, to determine what a model is actually using in its predictions — to better understand the technique, be able to explain how the model arrived at its decisions and therefore have more confidence in its performance. Consequently, we set out to investigate a facial feature extraction method that provides significantly more insight in what features it is using while making predictions for patients. We think this is particularly important in our clinical use case, since we are harnessing these techniques to diagnose vulnerable children.

All in all, this paper explores two new facial recognition technologies for classifying individuals with a genetic disorder and demonstrates improved performance compared to our previously published method [6]. We compare these methods on a dataset of 18 genetic syndromes and ensured that all algorithms used in this study are available on our GitHub repository.

## 2 Methods

### 2.1 Inclusion of individuals

We searched the literature for clinical studies, which included 2D facial photographs for 18 genetic syndromes associated with NDD. We ascertained that at least ten photographs per syndrome were available — otherwise, the syndrome was excluded from our dataset. The patients’ demographics in these studies and their pictures were collected and processed with our three facial feature extraction methods. When individuals were related, one individual of that family was chosen (based on the quality of the picture).

### 2.2 Data processing

Since we expected to have relatively few data available, 10-fold cross-validation was employed during the training and evaluation of the models. Furthermore, considering our dataset is imbalanced, both under- and oversampling were utilized in two separate settings. In one, all classes with less than 40 samples were randomly oversampled to 40. In comparison, all classes with more than 40 patients were randomly undersampled to 40 — so that every class contained 40 samples, regardless of the original number of patients. Since the sampling process is random, we iterated this process five times, leading in total to 50 folds (5 × 10). We did not add any more iterations, since our dataset is relatively small and therefore overlap in datasets would become inevitable, if the number of iterations was further increased.

Furthermore, in the second analysis, all classes with less than 20 samples were randomly over-sampled to 20. In comparison, all classes with more than 20 samples were randomly undersampled to 20 — ensuring that every class contained 20 samples. Again, this process was iterated five times — and in all five times, we used 10-fold cross-validation.

Of course, random oversampling can lead to overfitting on the minority class and therefore inflated performance results. However, we are more interested in the difference in classification performance between the different feature extraction methods and not so much in the absolute accuracy. Therefore, it is a valid approach in our use case.

### 2.3 Confounding analysis

In our primary analysis, all patients were included. However, there could be slight differences in possible confounding features of patients that could influence our predictions, like age, gender or ethnicity. Therefore, we created a subset of the data in which we included 15 patients of the five syndromes with the most patients. We matched these patients on gender, ethnicity, and age (measured in years). If a perfect age match was not available, we ensured age was within 1/3 of the age of the other patients. This resulted in a subset of the data of 75 patients of 5 different syndromes, matched on age-, gender- and ethnicity.

### 2.4 Extraction of facial features

To select the prime approach to extract features from a facial photograph, we investigated three methods: the previously published hybrid model [6, 9], the MediaPipe library [11] and a deep neural network: VGGFace2 [12, 13]. The best-performing method was ultimately incorporated in our final model (see Figure 1 for an overview of the methodology of this study).

**Figure 1:**
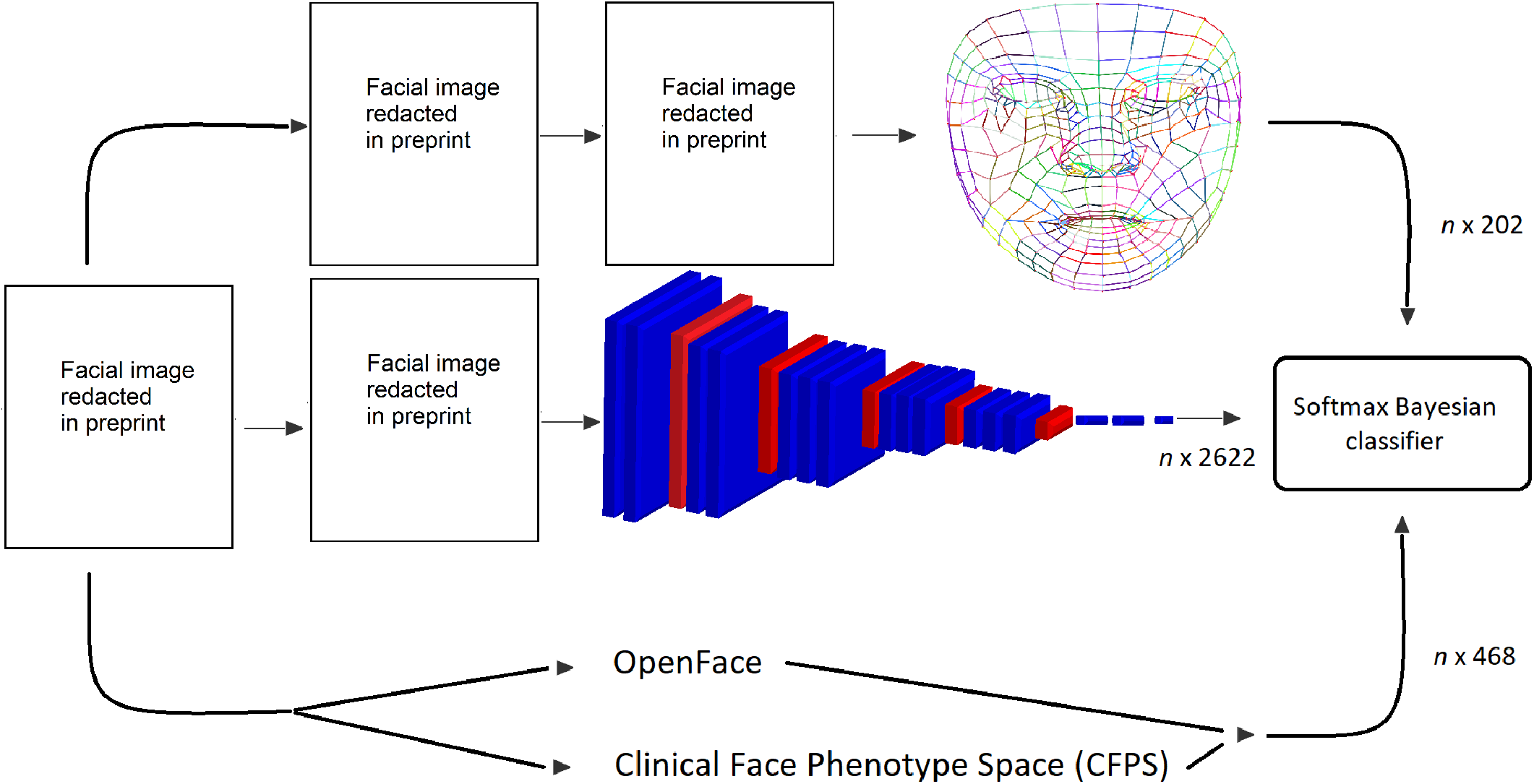
The global workflow of this study with the three facial features extraction methods displayed. Above, the MediaPipe pipeline: a face is first detected, then the 468 facial landmarks are detected, and these are then used to construct the face graph, consisting of 202 features, based on 1365 euclidean distances (edges with the same color belong to the same feature). The VGGFace2 pipeline is illustrated in the middle. First, face alignment is performed using MTCNN, after which the feature representation of the image according to VGGFace2 is obtained. In blue are the convolutional layers of the VGGFace2 architecture, including ReLu, while in red, the max-pooling layers are displayed. Finally, in the bottom row, the pipeline of the hybrid model is shown. The image is processed both by the CFPS and OpenFace, after which the feature vectors are concatenated. Photos used after written informed consent was obtained.

#### 2.4.1 Hybrid model

The hybrid model [6] makes use of two computer vision algorithms: the Clinical Face Phenotype Space (CFPS) [4] (trained on faces of patients with a genetic syndrome) and OpenFace [2] (a generic facial recognition algorithm trained on faces of the healthy population).

The CFPS pipeline uses facial landmark detection to obtain a face representation. This face representation constitutes of a 340-dimensional feature vector. On the other hand, OpenFace can be used to obtain a 128-dimensional feature vector of an image, using the pretrained weights of its deep neural network. For both representations, the idea is that people with similar faces have a similar feature vector, while dissimilar faces are further apart in the feature space.

Since the feature vectors are of different lengths but equally important, they were first normalized (L2 norm) before concatenation, assuming euclidean distance between data points. In the end, the hybrid model, therefore provides a 468-dimensional feature vector (340 from CFPS, 128 from OpenFace).

#### 2.4.2 MediaPipe

For the MediaPipe library, we first used a pretrained neural network to detect a face in a picture [11]. The picture was then cropped to the outline of the face, and the image was resized, preserving the aspect ratio. We then detected 468 facial landmarks for all individuals, and from those, a face mesh of 202 edges between these facial landmarks was created. These 202 edges were based on the original face graph of MediaPipe, which consists of 1365 edges between the 468 landmarks. However, this graph was manually amended since there was high collinearity between the facial distances. The length of the edges (corresponding to the Euclidean distance between the selected landmarks) was normalized (L2 norm). This approach has one advantage over the other two techniques: since it does not rely on a CNN but uses facial distances instead, we can easily visualize this pipeline’s features in its predictions, which greatly improves the explainability of this method compared to the other two.

#### 2.4.3 VGGFace2

Utilising VGGFace2 [12, 14, 13]; first MTCNN [15] was used to detect a face and crop the picture to the outline of the face. The feature representation of all cropped facial photographs was then obtained by using the pretrained deep neural network to process the images: the last layer before the classification layer was inspected to gather the feature vector according to VGGFace2.

### 2.5 Construction of probabilistic classifier

These feature vectors were used to train a softmax classifier. Considering we have relatively few data available per genetic syndrome (since these are rare), we preferred a Bayesian approach — since Bayesian inference allows the incorporation of prior knowledge in modelling. Therefore, the produced models tend to work better in small datasets. Furthermore, since we expected only a few features to be relevant (not all calculated distances between the facial landmarks will be important), we chose a softmax regression model with a feature-selecting prior.

More specifically, we implemented the logit-normal continuous analogue of the spike-and-slab (LN-CASS) prior [16] for the regression coefficients. Since it is continuous, inference is significantly easier than when using the original discrete spike-and-slab prior. Furthermore, the LN-CASS has shown improved performance when compared to other regularisation alternatives, such as the Finnish horseshoe [17], and can be considered the state-of-the-art in feature selection in Bayesian regression. All models were implemented in PyMC3 3.9 using the NUTS Hamiltonian Monte Carlo algorithm [18, 19]. For the mathematical definition of the model, please see the supplemental data.

## 3 Results

### 3.1 Study population

We included 524 non-familial individuals diagnosed with one of 18 different genetic syndromes in this study. In total, data from 68 different publications were used (see Table 1 for the complete overview of the demographics per genetic syndrome and Supplemental Table 1 for all publications used as sources for the data used in this study). Not all facial photographs were successfully processed, mainly due to misalignment of the facial landmarks: the hybrid model processed 452 pictures (86.3%), VGGFace2 all but one (99.8%), while the MediaPipe model correctly identified the face and corresponding landmarks in all (100%) of the cases (Table 2).

**Table 1:**
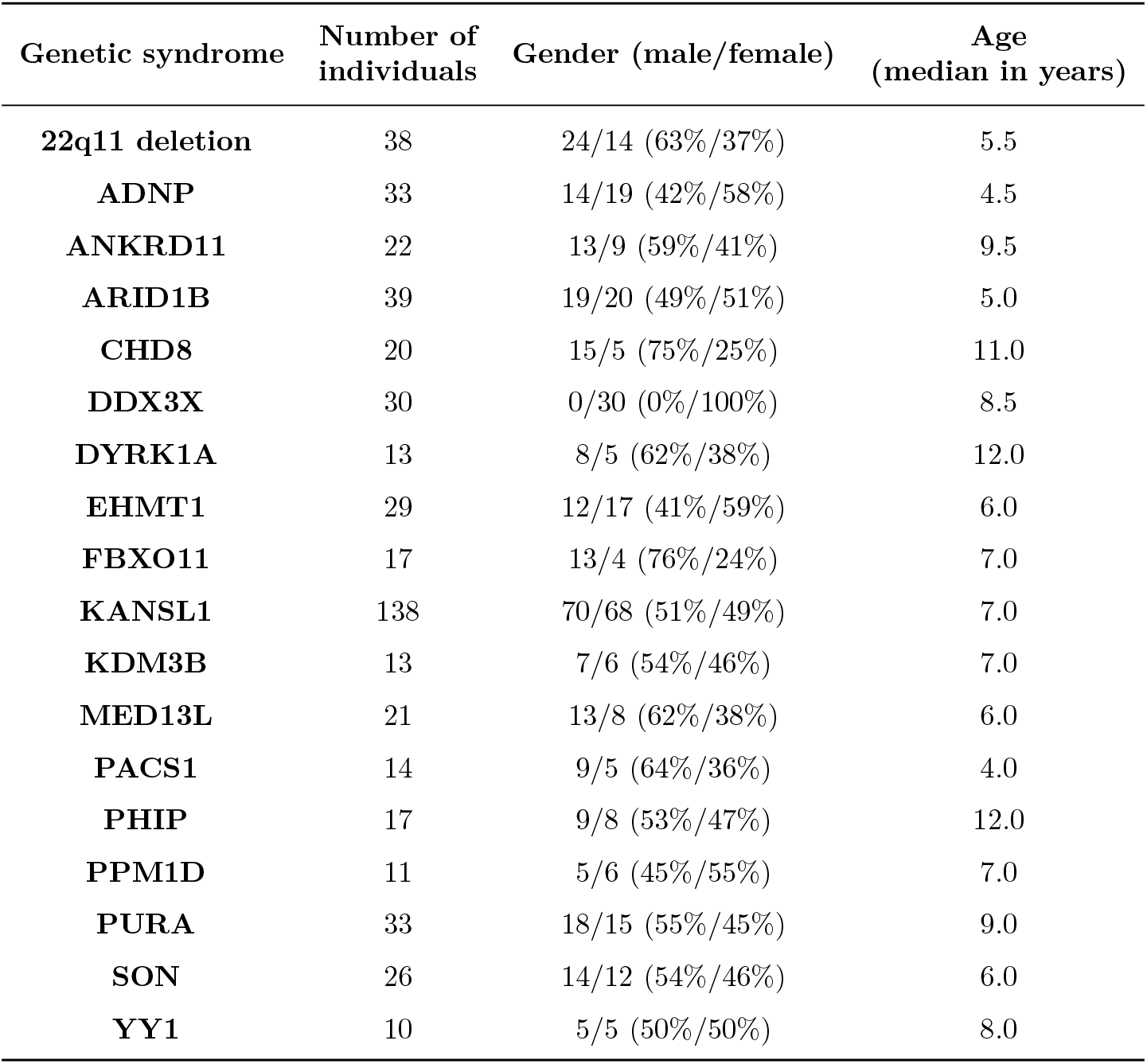
The number of individuals with a facial photograph per genetic syndrome available is shown here. Per genetic syndrome, the number of individuals selected, the gender distribution,and the median age are displayed.

| Genetic syndrome | Number of individuals | Gender (male/female) | Age (median in years) |
| --- | --- | --- | --- |
| **22q11 deletion** | 38 | 24/14 (63%/37%) | 5.5 |
| **ADNP** | 33 | 14/19 (42%/58%) | 4.5 |
| **ANKRD11** | 22 | 13/9 (59%/41%) | 9.5 |
| **ARID1B** | 39 | 19/20 (49%/51%) | 5.0 |
| **CHD8** | 20 | 15/5 (75%/25%) | 11.0 |
| **DDX3X** | 30 | 0/30 (0%/100%) | 8.5 |
| **DYRK1A** | 13 | 8/5 (62%/38%) | 12.0 |
| **EHMT1** | 29 | 12/17 (41%/59%) | 6.0 |
| **FBXO11** | 17 | 13/4 (76%/24%) | 7.0 |
| **KANSL1** | 138 | 70/68 (51%/49%) | 7.0 |
| **KDM3B** | 13 | 7/6 (54%/46%) | 7.0 |
| **MED13L** | 21 | 13/8 (62%/38%) | 6.0 |
| **PACS1** | 14 | 9/5 (64%/36%) | 4.0 |
| **PHIP** | 17 | 9/8 (53%/47%) | 12.0 |
| **PPM1D** | 11 | 5/6 (45%/55%) | 7.0 |
| **PURA** | 33 | 18/15 (55%/45%) | 9.0 |
| **SON** | 26 | 14/12 (54%/46%) | 6.0 |
| **YY1** | 10 | 5/5 (50%/50%) | 8.0 |

**Table 2:** The results of the three facial feature extraction methods after the three seperate analyses on the dataset are displayed here. First, the results of the analysis in which the data were randomly over- and undersampled to 20 per class are demonstrated, then to 40 per class, and finally the matched dataset. This dataset included 75 patients of 5 different syndromes (15 per syndrome), who were matched on age, gender and ethnicity, to correct for these possible confounders. *=statistically significantly better than MediaPipe; ** =statistically significantly better than MediaPipe and the Hybrid model.

|  | MediaPipe | VGG-Face | Hybrid model |
| --- | --- | --- | --- |
| **Accuracy (n = 20 per class)** | 0.18 [0.16-0.2] | 0.49 [0.46-0.52]* | 0.45 [0.42-0.48]* |
| **Accuracy (n = 40 per class)** | 0.34 [0.33-0.36] | 0.78 [0.76-0.8]* | 0.78 [0.75-0.81]* |
| **Accuracy (matched dataset)** | 0.29 [0.19-0.38] | 0.6 [0.5-0.71]* | 0.48 [0.27-0.68]* |
| **Log loss (n = 20 per class)** | 2.67 [2.63-2.71] | 1.89 [1.81-1.98]** | 2.02 [1.96-2.09]* |
| **Log loss (n = 40 per class)** | 2.25 [2.21-2.28] | 0.79 [0.71-0.88]** | 0.96 [0.85-1.08]* |
| **Log loss (matched dataset)** | 1.59 [1.46-1.71] | 1.15 [1.0-1.31]** | 1.4 [1.25-1.54]* |
| **Execution time (seconds per photo)** | 0.3 | 1.9 | 158.9 |
| **Succesfully processed photos (%)** | 100.0 | 99.8 | 86.3 |

### 3.2 Performance of facial feature extraction models

The two methods that depend on CNNs, the Hybrid model and VGGFace2, performed significantly better than the MediaPipe approach (Table 2). If we are looking at the over/undersampled dataset with 20 individuals per class, the accuracies of the three models are 0.18 for MediaPipe, 0.48 for VGGFace2, and 0.45 for the Hybrid model. When we increased the number per class to 40 by upsampling, the accuracy of all models improved — and the log loss was significantly reduced. The classification performance was comparable for VGGFace2 and the Hybrid model (0.78 in both cases), while the log loss was significantly lower when using VGGFace2. These results corresponded with the classification performance on the age-, gender- and ethnicity-matched dataset (Table 2).

Looking at the confusion matrices of VGGFace2, for instance (Figure 2), it was clear that some genetic syndromes were more challenging to predict than others — as would be expected. However, there was no clear imbalance in the predictions, and this was consistent across the three analyses.

**Figure 2:**
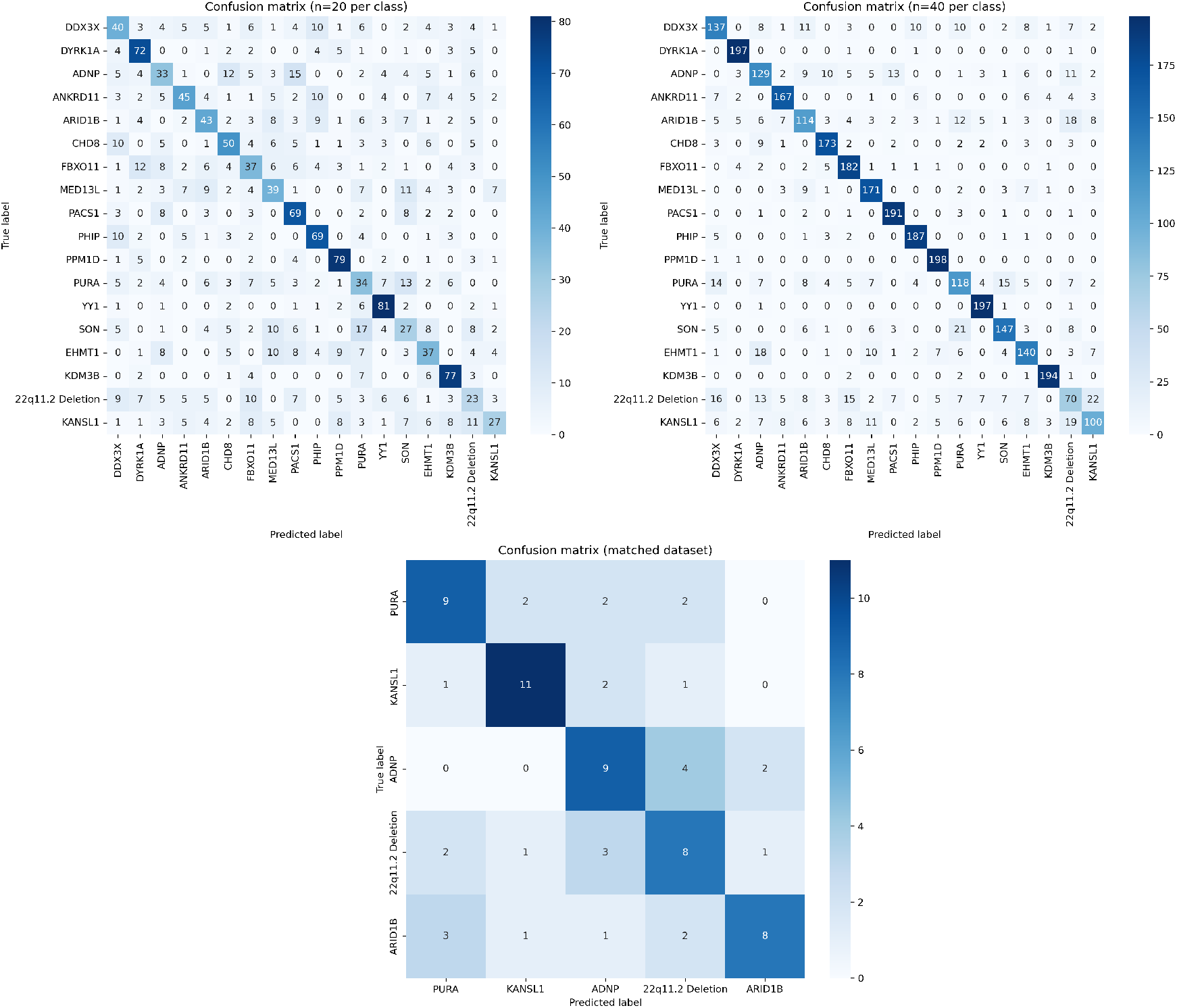
The confusion matrices of the predictions of VGGFace2 in the three performed analyses. While there are genetic syndromes that are harder to process for VGGFace2, overall, the predictions are quite evenly spread among the classes and reasonably consistent across the different investigations. Some classes, which are known to be less expressive in facial characteristics, are indeed harder to predict (*SON, FBXO11*). As expected, increasing the numbers per class (by oversampling) improves the performance.

Furthermore, the processing time of the older Hybrid model was significantly longer than that of the newer models: 158.9 seconds per photograph for the Hybrid model vs. 0.3 and 1.9 seconds per photograph for MediaPipe and VGGFace2, respectively.

### 3.3 Explainability of models

Due to the nature of the models, the MediaPipe method has significantly stronger explainability than the other two, since it relies on facial distances and not on a CNN, which are harder to inspect by nature. However, it is possible to create activation or feature maps for these CNNs. Activation maps are a visual representation of the activity of the hidden neurons of the network at a specific layer of a CNN. By visualizing these, we can obtain information on what parts of the input these neural networks are using while processing a particular image. Since VGGFace2 showed the best classification performance, we further investigated this model using this approach. Therefore, activation maps for VGGFace2 were generated (these figures are not available in the preprint because of the recognizable facial images). Using these maps, we show that VGGFace2 recognizes specific facial features, breaking these down into smaller and more abstract parts — until these are not recognizable features anymore.

#### 3.3.1 Local Interpretable Model-agnostic Explanations

To see if we could further interpret the face embeddings generated by VGGFace2, we used Local Interpretable Model-agnostic Explanations (LIME) [20, 21]. The main idea of this method is to train a local surrogate model to approximate the predictions of the model of interest. This local surrogate model is usually a relatively simple linear model — to be easily inspected and explained. By then perturbating original input data and inspecting the corresponding predictions, the importance of features can be shown. The power of LIME is that the model of interest can be any type of model, including black-box models such as CNNs.

For image data, the image is first segmented into several parts. These subparts of an image are called superpixels and these are then perturbed. By changing the images and inspecting the corresponding predictions, we can use LIME to weigh the relative importance of each superpixel. Since we are most interested in different facial parts, we used MediaPipe to segment each face into 11 different parts. These were then used to perturbate the images and train a Lasso regression model. The weights of the superpixels (and a superpixel corresponds to a complete facial part in our analysis) were then used to generate heatmaps, to visualize the most important facial regions for each genetic syndrome.

We performed this analysis for two different datasets: the full dataset of the 18 syndromes randomly over/undersampled to 20 individuals per class and the matched dataset, in which 15 individuals per syndrome of 5 syndromes were selected. For the full dataset, we generated the LIME during cross-validation, while for the matched dataset, we used the remaining individuals of those 5 syndromes, not in the matched dataset. The heatmaps were then generated by looking at the top 5 (for the matched dataset, since it is not oversampled and therefore smaller) and top 10 (for the over/undersampled dataset) correctly predicted individuals per syndrome. The weights per superpixel were averaged, as to correct for any outliers and to provide a reliable assessment of the importance of each facial part.

When looking at the matched dataset (Figure 3), the most important features per syndrome are the forehead for *PURA*, the nose for *KANLS1*, the forehead for *ADNP*, quite evenly spread for the individuals with the 22q11.2 deletion syndrome, and the nose for *ARID1B*. Especially the forehead for *ADNP* and the nose for *KANSL1* correspond remarkably well with the facial parts described as most prominently different and important in these specific syndromes by experts. In the larger, over/undersampled dataset (Supplemental Figure 1), interestingly, for *KANSL1*, the nose is no longer the most important feature. However, classification performance is significantly worse than in the matched dataset, as indicated by both the lower positive predictive value and the lower sensitivity. Therefore, this unexpected behaviour is probably due to the task not being learned sufficiently well in the first place. In this analysis, however, *PPM1D* shows remarkable predictive performance and in this case, the mouth area is prioritized as an important feature. Again, this agrees with the dysmorphic features deemed important by clinicians when assessing this genetic syndrome.

**Figure 3:**
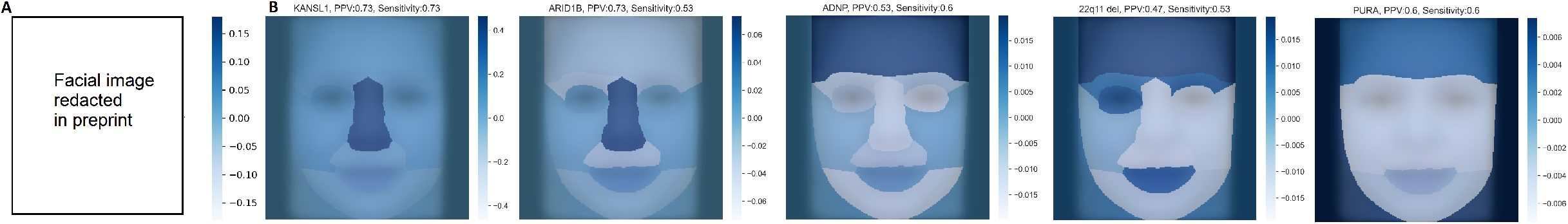
A) The most important facial parts in the prediction of a single individual with *KANSL1*, using LIME with the classifier trained on the matched dataset. Here, darker shades of blue indicate more important parts according to LIME. The nose is clearly the most important feature in this case, which corresponds to our intuition in *KANSL1*. B) The generated heatmaps using LIME of the five genetic syndromes in the matched dataset. Per genetic syndrome, the weights of the different facial parts were average over the top five correctly predicted individuals per syndrome. The averaged weights were then visualised in a heatmap. To provide an idea of the predictive performance per syndrome, the positive predictive value (PPV) and sensitivity are shown, as calculated from the same data as the heatmaps in Figure 2. Some syndromes, like *KANSL1, ADNP* and *ARID1B*, have more pronounced important features. Others are more evenly divided, like *PURA* and the invididuals with 22q11.2 deletion syndrome.

## 4 Discussion

In this study, we explored three facial feature extraction methods in recognizing 18 genetic syndromes in 524 individuals. We show that the approaches that rely on convolutional neural networks (CNNs), VGGFace2 and the Hybrid model, display superior classification performance compared to a facial landmark approach. Furthermore, VGGFace2 processes facial images significantly faster than the Hybrid model (in seconds instead of minutes) and seldom fails to process an image. Both these properties are important when aiming for clinical use, since a clinician will not use a method that needs minutes to process an image, or often fails to process an image at all. Consequently, we are of the opinion that VGGFace2 is currently the best open-source methodology available to extract facial features when working with individuals with dysmorphic faces.

Others have previously investigated using computer vision techniques on facial photographs of children with suspected genetic syndromes, for instance to demonstrate that children with cancer at a young age might be facially different from controls [22]. Looking at rare genetic disorders that cause neurodevelopmental disorders, classifying individuals using deep learning has been successfully applied as well [4, 5, 6, 7, 8, 9, 10] — some with significantly larger datasets than we present here. However, these studies use proprietary algorithms unavailable to the academic community. Validating, confirming, or building on these algorithms is therefore problematic. In contrast, we are open-sourcing all code used in this study so that others can further improve on our methods. Furthermore, since these other studies are retraining their CNNs, the risk of introducing biases and overfitting increases.

While we confirm CNNs are powerful, one of the distinctive properties of these networks is that these do not provide insight on what basis they are generating their output. Indeed, a famous example of results that appear impressive at first is the classification of possible melanomas using these CNNs. Although the authors noted remarkable and dermatologist-level results at first, later studies showed that the predictions of these CNNs were influenced by a ruler in the image of a skin lesion — since the presence of a ruler was associated with malignancy [23, 24, 25].

Returning to our use case, using a CNN in an end-to-end manner to assist a clinician in diagnosing a patient based on a facial photograph does not shed light on what the CNN is using to make its predictions. Is it looking at dysmorphisms, like we do? Or is it picking up biases in the data sets — like emotional expressions or particular preferences of a specific clinician taking a photograph? We circumvent this problem by not training a new CNN or retraining VGGFace2, but using the pretrained weights, learned in the general facial recognition task in the sizeable VGGFace2 dataset (3.1 million individuals) [12, 13]. In this manner, we ensure that we are using actual facial data in the classification and not other information in the photographs unrelated to facial recognition. Since VGGFace2 is unaware of the classification labels, it is impossible to overfit on that information or learn spurious correlations. We further verify the information VGGFace2 uses in the photographs on validity by generating activation maps (**??**, supplemental data). These confirm that we look at facial features and not other irrelevant information.

To further investigate VGGFace2, we used Local Interpretable Model-agnostic Explanations (LIME). By first segmenting the faces of individuals in different facial parts, perturbating the images and inspecting the corresponding changes in the predictions, we can both look at the importance of features in a single prediction, and visualize the most important facial parts per genetic syndrome as a whole (see Figure 3 and Supplemental Figure 1). When the predictive performance of a specific syndrome is satisfactory, the most important features per genetic syndrome correspond remarkably well to the features deemed important by clinicians. Examples of these are the mouth in *PPM1D*, the nose in *KANSL1* in the matched dataset, and the forehead in *ADNP* (in the matched dataset as well) [26, 27, 28]. When the predictive performance is not as good, as for instance for *KANSL1* in the under/oversampled dataset, the important features are not recovered as well anymore. It shows that clinicians correctly deem these particular features important in recognizing these disorders — since when we do not correctly estimate the importance of these facial parts, predictive performance degrades. Furthermore, these findings confirm that VGGFace2 and our model consider facial parts that clinicians too deem meaningful in general facial recognition and individuals with dysmorphic faces. This is important, as it shows our model is considering semantically meaningful facial parts, further strengthening our confidence in this method for clinical use.

A limitation of this study (and of using artificial intelligence in clinical genetics in general) is the use of a relatively small and imbalanced dataset. This is inevitable since the described genetic syndromes are rare diseases that are by definition uncommon and small in affected individuals. We attempted to correct for the imbalance in the dataset, by random over- and undersampling. Certainly, random oversampling can lead to overfitting the model on the minority class. However, we emphasize that in this study, we are not interested in the absolute classification performance of the models, but in the relative performance of the facial feature extraction methods when compared to each other — to select the best technique. Any possible inflation of classification performance is equal for all three methods, and therefore this approach is well-suited for our aim (evaluating the best technique out of these three).

### 4.1 Conclusion

The classifier using the features extracted by VGGFace2 not only shows superior classification performance but detects faces in almost all images that are processed. Furthermore, processing images takes seconds, making it feasible to use in a clinical setting. By not retraining VGGFace2, but instead using the feature vector of the network with its pretrained weights, we avoid overfitting our model. We offer insight into the important features according to VGGFace2 and show that these features correspond to facial parts experts prioritize when recognizing these genetic disorders. We confirm that it is possible to classify individuals with a genetic disorder using artificial intelligence and open-source all of the models used in this study.

## Data Availability

The machine learning model and code created during this study are freely available at
https://github.com/ldingemans/FacialFeatureExtraction. The used dataset is not publicly avail-
able due to both IRB and General Data Protection Regulation (EU GDPR) restrictions, since the
data might be (partially) traceable. However, access to the data may be requested from the data
availability committee by contacting the corresponding author.

## 5 Acknowledgements

We are grateful to the Dutch Organisation for Health Research and Development: ZON-MW grants 912-12-109 (to B.B.A.d.V. and L.E.L.M.V.), Donders Junior researcher grant 2019 (B.B.A.d.V. and L.E.L.M.V.) and Aspasia grant 015.014.066 (to L.E.L.M.V.). The aims of this study contribute to the Solve-RD project (to L.E.L.M.V.) which has received funding from the European Union’s Horizon 2020 research and innovation programme under grant agreement No 779257. Furthermore, we would like to thank Jayne Y. Hehir Kwa for fruitfull discussions.

On the ethical side, in this study, data from the ‘Biobank Intellectual Disability’, which is part of the Radboud Biobank initiative (for more information, see [29] or https://www.radboudumc.nl/en/research/radboud-technology-centers/radboud-biobank) were used. Within this biobank, phenotypic and molecular data have been systematically captured for individuals with (non-)syndromic ID referred to the Radboud university medical center. The use of this dataset was approved by the ethical committee of the Radboud university medical center (#2020-6151). Furthermore, the authors declare no competing interests.

## 5.1 Data and code availability

The machine learning model and code created during this study are freely available at https://github.com/ldingemans/FacialFeatureExtraction. The used dataset is not publicly available due to both IRB and General Data Protection Regulation (EU GDPR) restrictions, since the data might be (partially) traceable. However, access to the data may be requested from the data availability committee by contacting the corresponding author.

## 6 Supplementary data

### 6.1 Mathematical definition of Bayesian softmax regression model

Suppose dataset *Z* = (*X*_*i*_, *y*_*i*_) consists of *n* patients, where the *i*-th patient is represented by their *p* symptoms *x*_*i*_ = [*x*_1_, *x*_2_, *x*_3_, …, *x*_*p*_] (with *p* = 202, 2622 or 468 for the three different feature extraction methods) and *y*_*i*_ ∈ {0, 1, …, 18} is the corresponding genetic syndrome and therefore a categorical variable. Then our aim is to predict *y*_*i*_ given the input features *X*_*i*_. Since *y*_1_ is categorical, we chose a softmax regression model.

Looking at this model, we chose the logit-normal continuous analogue of the spike-and-slab (LN-CASS) prior [16] as the prior for the regression coefficients. In the original spike-and-slab, a Bernoulli prior is used for the inclusion probability. Since the Bernoulli is a discrete prior, inference is relatively hard. By transforming the prior for the inclusion probability in a logitnormal distribution, we preserve the advantages of an inclusion probability per feature, but make inference significantly easier.

Going more into detail about the LN-CASS prior, we select a normal distribution with mean zero as the prior for the regression coefficients. The variance of the distribution is defined by *τ* (the “unregulated” variance) and *λ*_*p*_ (inclusion probability of this feature). We chose a HalfNormal(2) prior for the unregulated variance — since it needs to be positive, and relatively small. The prior for the inclusion probability *λ*_*p*_ is defined as a normal distribution, with mean *µ*_*λ*_ and standard deviation 10. Larger standard deviations create a stronger prior, in which inclusion probabilities are more likely to become zero or one. We chose 10, following the recommendations of the authors of the original LN-CASS paper.

To transform this normal distribution in a logit-normal distribution, *µ*_*λ*_ is the logit of the inclusion probability. In our use case, we chose logit^*-*1^(0.1) — since we estimate the inclusion probability for each feature to be around 10%.

All in all, the mathematical definition is therefore:

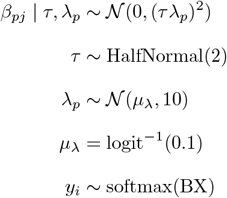

where B corresponds to all regression coefficients, and *X* to the feature vectors of size *n* × *p*

Convergence of the models was checked by sampling from two chains using PyMC3, checking the R-hat statistic and checking for divergences. Both chains were sampled for 2000 samples, after which 1000 were discarded after tuning.

**Supplementary figure 1:**
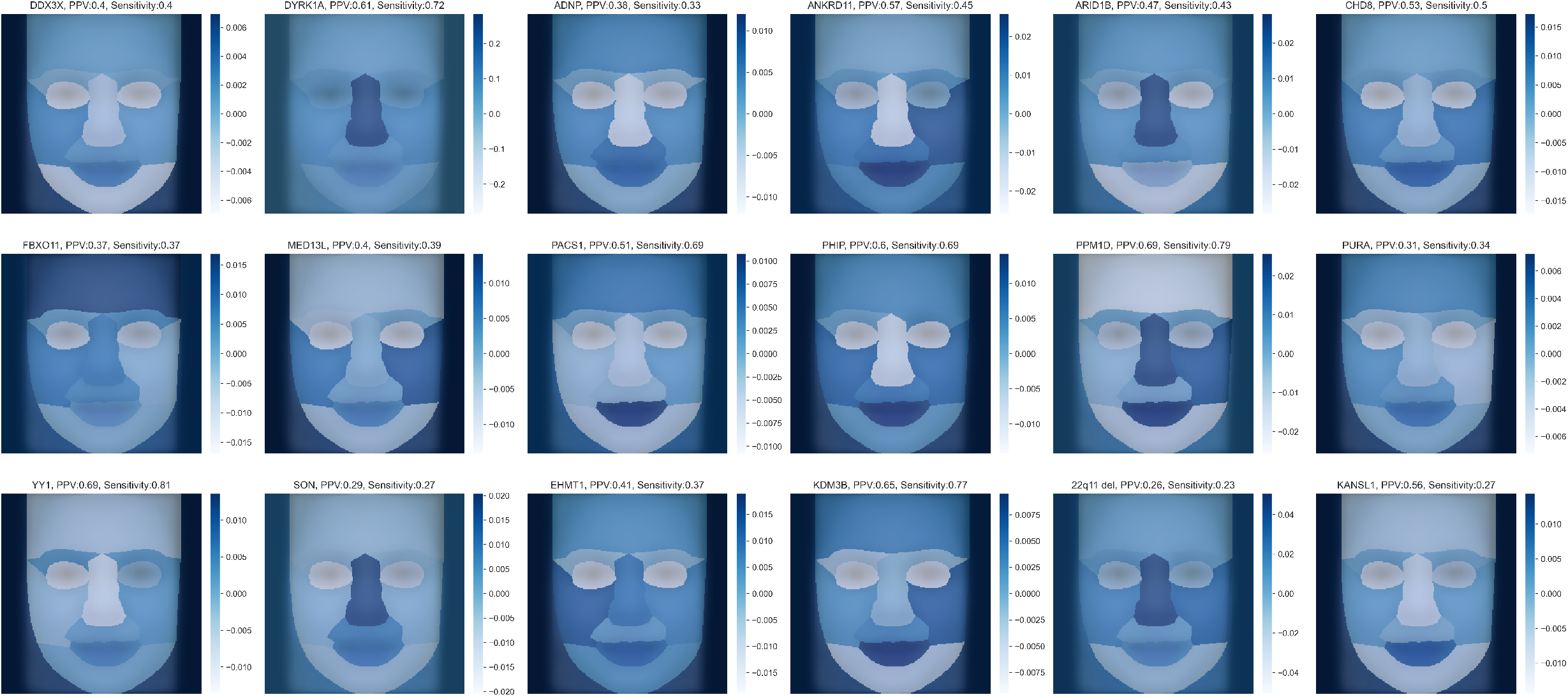
The generated heatmaps using LIME on the full dataset, over/undersampled to 20 individuals per genetic syndrome. The positive predictive value (PPV) and sensitivity are shown, as calculated from the same data as the heatmaps in Figure 2. Again, some genetic syndromes show particular important facial features, while others show a more even distribution of the weights per facial part.

**Supplementary table 1:** A list of the used publications per syndrome to create the dataset by extracting the phenotypic data and photographs of individuals in these papers. For several syndromes, not (yet) published individuals were added to the dataset as well, as indicated by Not published.

| Gene | PMID of used publications |
| --- | --- |
| **22q11 deletion** | 15831592,17041934,18636631,1956057,21200182,25317860,3816857,12548732 |
| **ADNP** | 24531329,25217958,27031564,29724491 |
| **ANKRD11** | 19920853,21527850,21654729,22307766,23494856,26269249 |
| **ARID1B** | 19034313,22405089,22426309,22585544,23906836,24569609,26395437,26754677,27112773,28323383,30055038,30349098,31981384,32339967 |
| **CHD8** | 23160955,24998929,31001818,31721432,Not published |
| **DDX3X** | 26235985 |
| **DYRK1A** | 18405873,21294719,23099646,23160955,25707398,Not published |
| **EHMT1** | 22670141,Not published |
| **FBXO11** | 27479843,28343630 |
| **KANSL1** | 16906164,17601928,18628315,21094706,22544363,26306646,Not published |
| **KDM3B** | 30929739 |
| **MED13L** | 23403903,24781760,25712080,28645799,29511999,29959045 |
| **PACS1** | 26842493 |
| **PHIP** | 29209020 |
| **PPM1D** | 28343630 |
| **PURA** | 27148565,29097605,29150892 |
| **SON** | 24896178,27256762,27545680,34521999 |
| **YY1** | 21076407,28575647 |

